# Modeling COVID-19 in Iran using Particle Swarm Optimization algorithm

**DOI:** 10.1101/2021.04.10.21255244

**Authors:** Ebrahim Sahafizadeh, MohammadAli Khajeian

**Affiliations:** Department of Computer Engineering, Persian Gulf University, Bushehr 75169, Iran

**Keywords:** SARS-CoV-2, Pandemic, Public health, isolation, quarantine

## Abstract

**Background:** The first confirmed cases of COVID-19 in Iran were reported on February 19, 2020. The coronavirus expanded rapidly in all Iranian provinces and three waves of COVID-19 cases have been observed since the pandemic took effect and the fourth wave of Covid-19 cases will likely be observed soon. This study aimed to model the spread of COVID-19 in Iran and to estimate the epidemic parameters and to predict the short-term future trend of COVID-19 in Iran.

**Methods:** We proposed a modified SEIR epidemic spreading model and we used data from February 20, 2020, to April 9, 2021, on the number of cases reported by Iranian governments to fit the proposed model on the reported data. Particle Swarm Optimization (PSO) algorithm was employed to estimate the parameters of the proposed model and the numerical simulation results were obtained by Runge-Kutta method. The estimated parameters were employed to calculate the effective reproduction number and to predict the short-term future trends of COVID-19 cases.

**Results:** The results indicated that the effective reproduction number has increased during Nowruz (Persian New Year) and it was estimated to be 1.28. Considering only two exposed cases as the initial cases in the model, the cumulative number of exposed cases was estimated to be 15,252,372 individuals since the beginning of the outbreak. The prediction of the short-term future trends of COVID-19 cases with different scenarios showed that another peak of the pandemic cases occurs in the next weeks. By immediate lockdown implementation the number of active infected cases was estimated to be 397,585.

**Conclusion:** Different scenarios of short-term prediction of the future trends of COVID-19 cases indicated that immediate strict social distancing policies need to be implemented to prevent a tremendous burden of the fourth major wave of COVID-19 infections on the health care system of Iran.

## 1. Background

Iran reported the first confirmed cases of COVID-19 on February 19, 2020. In response to rapid growth of the pandemic, the government enforced social distancing in the majority of provinces. Closure of schools and universities, cancelation of religious gathering and social events, closure of Business and offices and reduction of working hours, and travel restriction, along with necessary instructions and advice have been the various interventions and policies implemented by government to control the spread of COVID-19 in different periods. However, three peaks of COVID-19 pandemic have been observed since the pandemic took effect. Based on the official reported data, the number of daily cases has started to grow up recently, and there is a possibility of observing the fourth wave of COVID-19 cases.

In this paper, SEIR epidemic model [1] is adapted to study the dynamics of the COVID-19 spreading in Iran. For this purpose, we propose a modified SEIR model and introduce the dynamical equations of the proposed system. We then conduct numerical simulation using Runge-Kutta method in MATLAB under various epidemic parameters generated using PSO algorithm to fit on the reported data. The estimated parameters are then used to predict short-term trends of COVID-19 under different scenarios.

## 2. Methods

### A) Model

We introduce the proposed model using a set of ordinary differential equations. At each time, each individual is in one of the following states:

- Susceptible (S): the individuals who are vulnerable to disease.
- Exposed (E): the individuals who are infected and has potential to spread but not officially confirmed.
- Infected (I): the individuals who are officially confirmed to be infected by coronavirus.
- Removed1 (R1): the individuals who have been exposed but they have not been officially confirmed and they do not transmit virus anymore (i.e. A fraction of Exposed class who are either dead or recovered).
- Removed2 (R2): the individuals who have already been confirmed but they either have been recovered or dead.

Fig. 1 shows the state changing graph of the model. As shown in Fig. 1, a susceptible in a contact with an exposed might change to an exposed case with rate *β*. A proportion of exposed cases which later test positive for COVID-19 are considered as infected class. We suppose that the confirmed cases (Infected class) are either in the hospitals or isolated at home, hence their infectious rate is neglected.

**Fig. 1.**
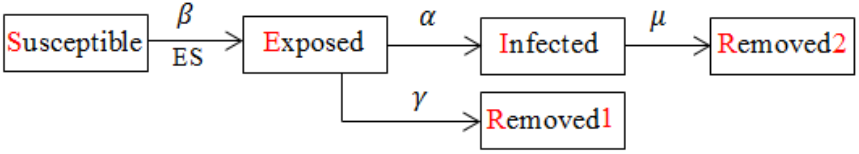
State changing graph of the model

When a susceptible person gets the disease, her/his state changes to exposed class in the model and transmits the virus to her/his contacts with rate *β*. A person in exposed class is either confirmed as an infected individual after *d* = 1/*α* days or cured and transmitted to Removed1 class after 1/*γ* days. The people in the infected class are supposed to be confirmed cases; hence they are isolated at home or in the hospital and we suppose their contact with susceptible people can be ignored in the model. We also considered *α* = *γ* in the model.

Let S(t), E(t), I(t), R_1_(t) and R_2_(t) be the number of susceptible, exposed, infected, removed1 (cases who have not been officially confirmed and do not transmit disease anymore) and removed2 (people who confirmed and cured or died) nodes at time t respectively. Suppose 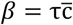 where τ is the probability of infection because of a contact between exposed and susceptible individuals and 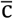 is the average rate of contact for each individual. So, each exposed person can infect *β* susceptible individuals per unit time. The infection rate can be reduced by reduction of infection probability (for example mandatory face mask in public) or by contact rate reduction (for example by schools and universities closure). The proposed model ODEs are as the following:

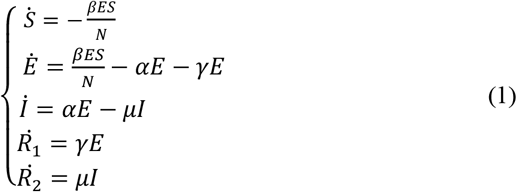

Where *N* is the population of Iran considered to be 83,992,949 [2].

### B) Reproduction number

The basic reproduction number, *R*_0_, is the average number of secondary cases generated by a single case during infectious period in a population where all individuals are susceptible to infection and there is no immunity in the population. The effective reproduction number, R_t_, however, is defined as the average number of secondary cases per infectious case when there is some immunity in the population and it can be estimated by the product of *R*_0_ and the fraction of the host population.

Considering the proposed model, the change of the number of exposed individuals is:

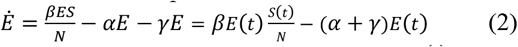

In a complete susceptible population 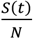 can be approximated by 1, and then the basic reproduction number of the proposed model can be calculated as the following:

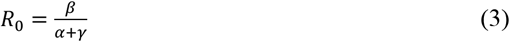

We used the same formula to calculate the effective reproduction number in different periods of time.

### C) Data

We use the official reported data from February 20, 2020, to April 9, 2021, reported by MoHME [3]. The total number of deaths plus the total number of recovered is considered as the number “Removed2”, and active confirmed cases are considered as “Infected” in the model at each time step.

### D) Error estimation

The fitting of the proposed model to the data is done in MATLAB by minimizing the root mean square error (RMSE) as the following:

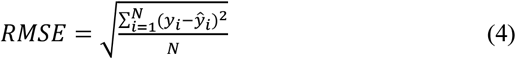

We used the daily reported data as the values of *y* and model generated data as the values of *ŷ* to calculate the error. This method provides the value of cost function for PSO algorithm to evaluate the set of model parameters that provide the best fit of model to the data.

### E) PSO algorithm

PSO is a population-based optimization technique originally proposed by Keneddy and Eberhart [4]. This algorithm is inspired from intelligent collective behavior of some animals such as flocks of birds where these swarms search for food in a collaborative manner. The goal of an optimization problem is to determine a variable represented by a vector *X*=[*x*_1_*x*_2_*x*_3_…*x*_*n*_]to optimize (minimizes or maximizes) an objective function *f*(*X*).

We use PSO algorithm to estimate the parameters of the proposed model to provide the best fit of the model to reported data. At each iteration, the parameters are estimated by PSO and they are passed to a function to solve ODEs. The Runge-Kutta method is used to solve the ODEs and then the error is calculated according to Eq (4). This error is considered as the cost function of PSO for the next iteration. The process continues to provide the best fit.

At the beginning of the process, all individuals are susceptible except individuals who are exposed at the initial state. We suppose the number of initial exposed cases to be two cases. The initial state is considered 10 days before the date that the first cases reported by the government. We run PSO algorithm to estimate the epidemic parameters such that the number of infected cases in the model equals to the number of first reported cases. Then we consider Feb 19, 2020 as *t*_0_ and the previous estimated exposed number as E(0). The number of the first reported cases was used as the initial numbers of each classes. Then we run PSO algorithm again and repeat the same process to fit the model on the reported data in different time period and estimate epidemic parameters for different time periods. Note that the estimated number of *E* at the end of each period is used as the initial number of *E* at the next period.

### F) Prediction

We employ the estimated values of the fitted parameters to predict the short-term future trends of COVID-19. These values are estimated for different previous periods of time including the period where strict social distancing implemented by the government, the period where the cases highly grew up, the period before Nowruz and the period during Nowruz.

## 3. Results

Fig 2. shows the curves of reported data and model generated curves in different periods. As can be seen in Fig.2, the model generated data successively fit on the reported data. The estimated parameter values of the model and the error of the fit are shown in Table 1.

**Table 1.**
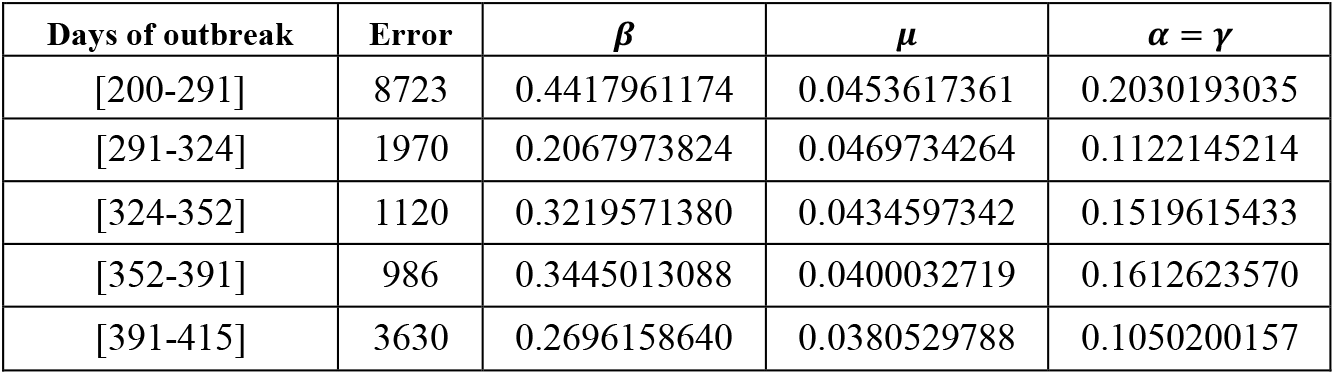
The estimated parameters

**Fig 2.**
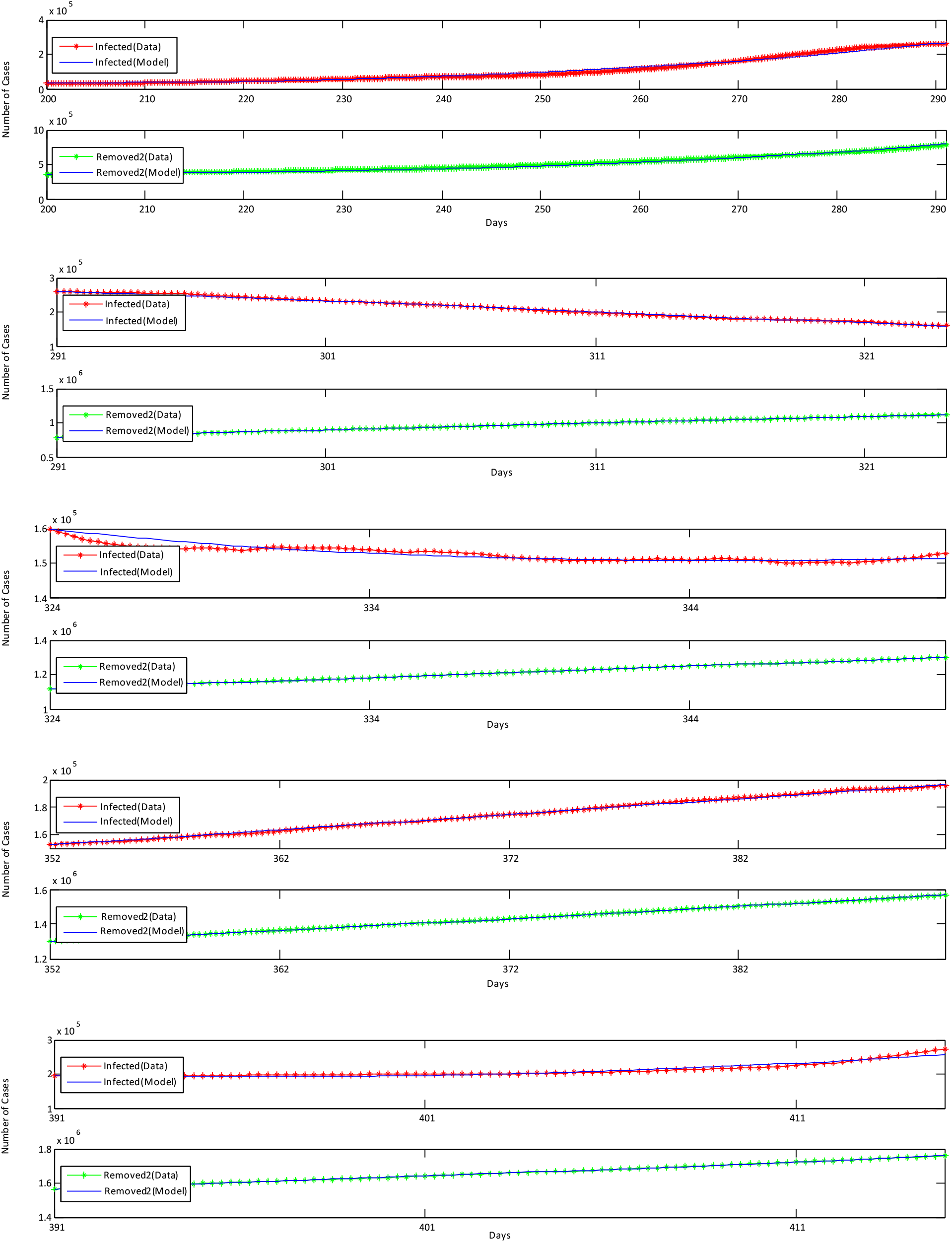
The curves of reported data and model generated data in different periods

According to Eq (3) and the estimated parameters in Table.1 we calculate the effective reproduction number in the different periods. Fig. 3 illustrates the curve of effective reproduction number. As can be seen in Fig. 3, the effective reproduction number has increased during Newruz and it is estimated to be 1.28. The curve of effective reproduction number illustrates that it is consistent with the curves estimated by JRC [5] and RKI [6] methods in the previous work [7].

**Fig. 3.**
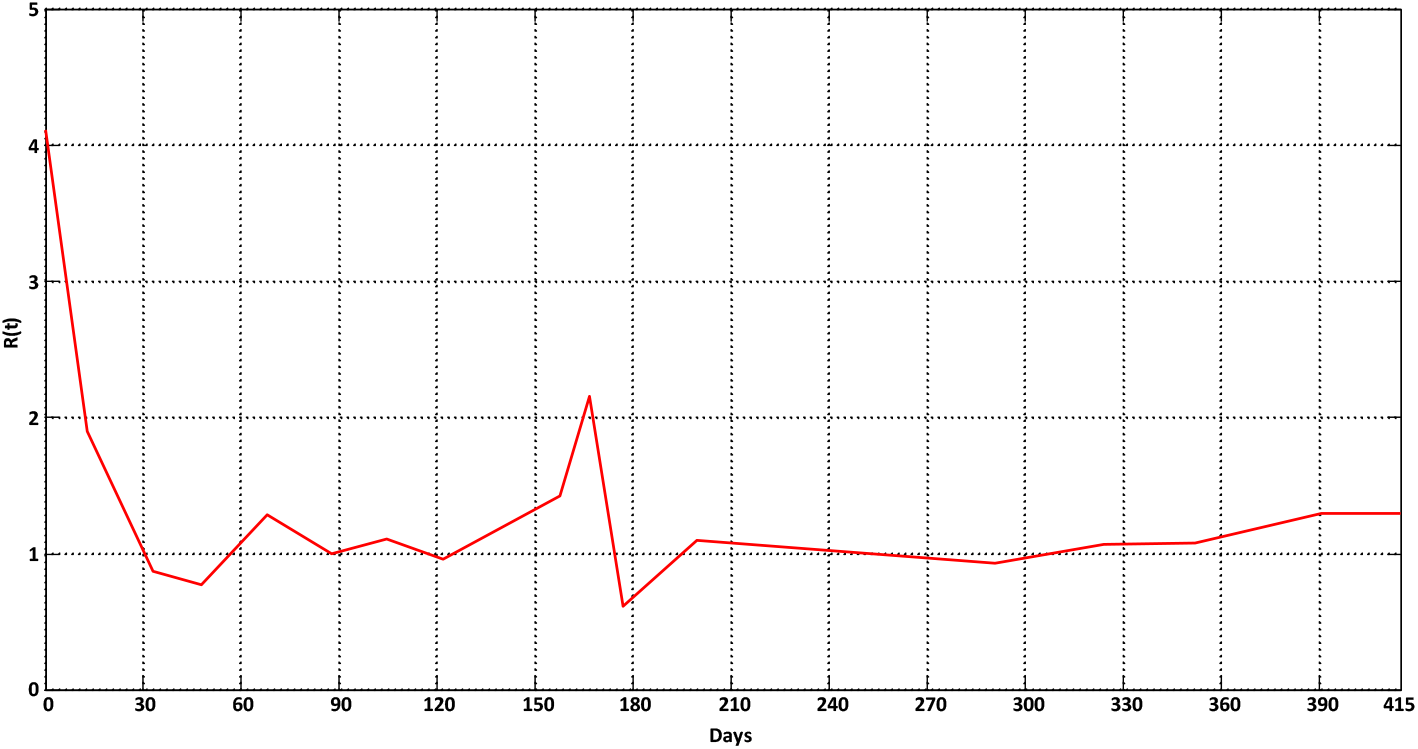
The curve of effective reproduction number

The estimated values of the fitted parameters are employed to predict the short-term future trends of COVID-19 in Iran. We begin with 415th day of the outbreak that we successively fitted the model to its previous days. The Estimated value of active exposed cases is used as the initial exposed cases and the last reported data is used to initialize the other parameters. Then we use different parameters of the model estimated by PSO algorithm in the previous periods of time to estimate the values for the cases in the next days. Table. 2 shows the parameters estimated in the period used for different scenarios as the following:

**Table 2.**
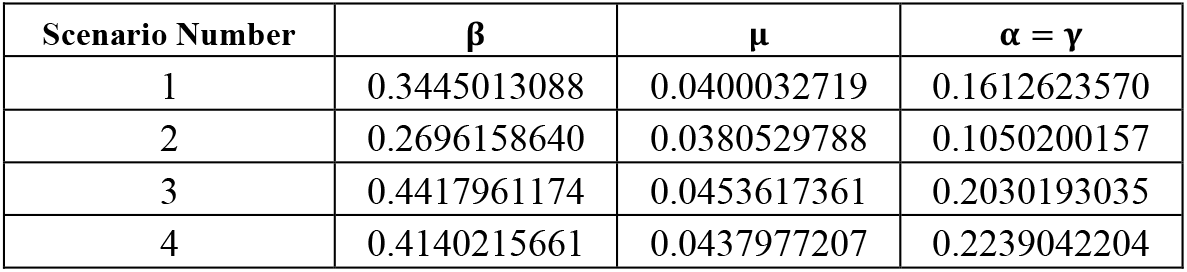
Epidemic parameters used for prediction

| Scenario Number | $\beta$ | $\mu$ | $\alpha = \gamma$ |
| --- | --- | --- | --- |
| 1 | 0.3445013088 | 0.0400032719 | 0.1612623570 |
| 2 | 0.2696158640 | 0.0380529788 | 0.1050200157 |
| 3 | 0.4417961174 | 0.0453617361 | 0.2030193035 |
| 4 | 0.4140215661 | 0.0437977207 | 0.2239042204 |

Scenario 1: Includes the parameters of period before Nowruz (days 352 to 391). These parameters are used to show the trend from 415^th^ day if the policies of that period would have been continued in Nowruz.

Scenario 2: Includes the parameters of the period during Nowruz (days 391 to 415).

Scenario 3: includes the parameters of the period in which the cases highly grow up (days 200 to 291).

Scenario 4: includes the parameters of the period in which the government implemented strict social distancing to control the rapid growths of new cases (days 282 to 305).

Fig. 4 shows the prediction of the number of infected cases (active confirmed case) for next five weeks with different scenarios.

**Fig. 4.**
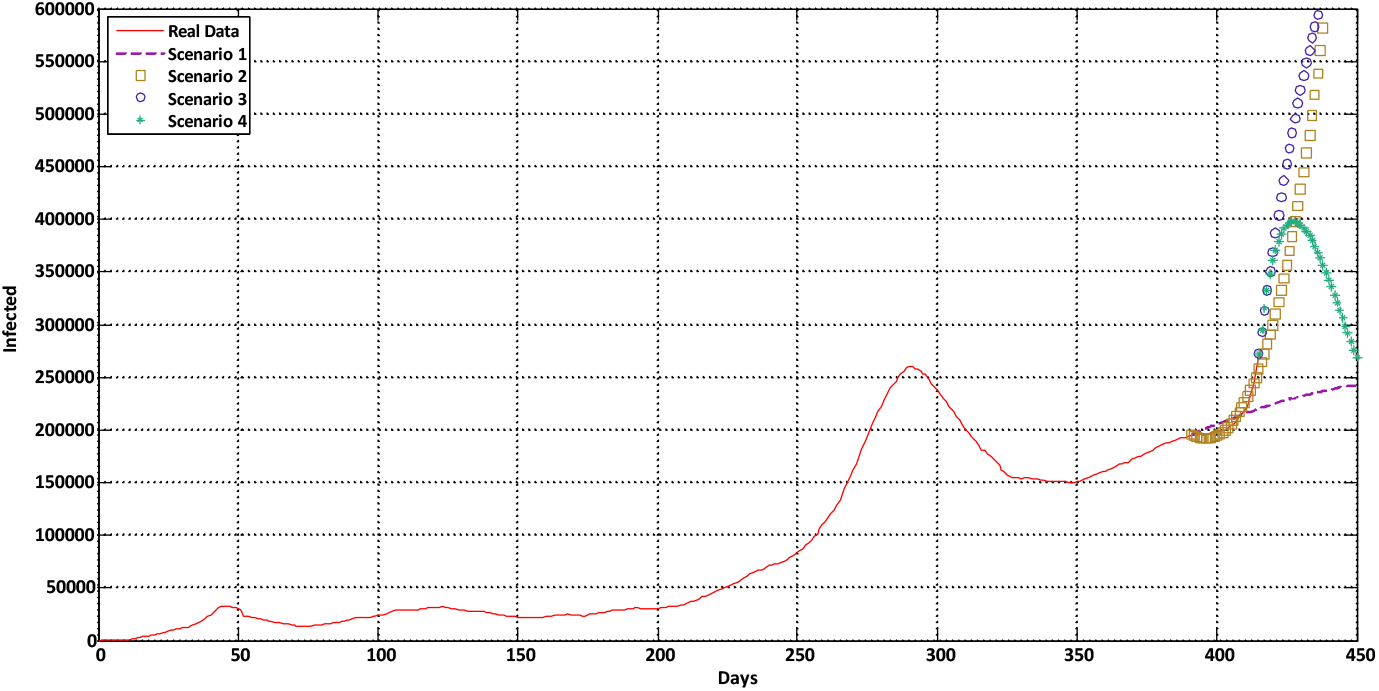
The future trend of epidemic

## 4. Discussion

Various researchers in different fields have carried out researches on COVID-19 in Iran [8][9][10][7]. Some studies have tried to investigate the relationship between COVID-19 and meteorological and climatological factors [11][12]. Several researches have employed mathematical models to analyze the epidemic curve and forecast the epidemic trend of COVID-19 Iran [13][14][15]. The main goal of the modeling of COVID-19 is to estimate the parameters to predict the future trends of the epidemic which is a valuable result to control the epidemic.

We proposed a modified SEIR epidemic spreading model and we used data from February 20, 2020, to April 9, 2021, on the number of cases reported by Iranian governments to fit the proposed model on the reported data. We employed PSO algorithm to estimate the parameters of the proposed model. Furthermore, we used the reported data along with the estimated parameters of the model to forecast the trends in the next weeks.

The prediction results in Fig. 4. illustrate different trends with different scenarios. Scenario 1 shows the trend with policies before Nowruz. However, Iranian people traditionally travel during Nowruz holidays and visit family and friends. Fig. 4 indicates that the effective reproduction number has been increased during Nowruz. Iran has already faced three major waves since the pandemic took effect. Based on the official reported data, the number of daily cases has started to grow up after Nowruz holidays, and there is a probability for the fourth wave of COVID-19 cases. Scenario 2 shows the trend with the parameters of Nowruz without any control and Scenario 3 shows the trend with the parameter of the third wave which the number of infected will quickly grows up in both of them. Scenario 4 shows the trend with parameters of the period in which the government implemented strict social distancing to control the third wave. If a strict social distancing is implemented immediately the number of the fourth peak is estimated to be 397,585.

There are some limitations in our study. First, the proposed model was set up based on a number of necessary assumptions. For example, we assumed that officially confirmed cases do not spread the disease after confirmation due to their contact limitations; we also assumed that the duration to be confirmed or transmitting to removed class due to self-quarantine are equal for exposed class. Second, the accuracy of the estimated exposed cases depends on the initial parameters such as the number of initially exposed cases which is a limitation. And finally, in term of prediction, our model uses the previous estimated parameters, assuming that the previous patterns continue in the future. However, the authorities often change their control strategies and the actual parameters significantly depend on these strategies which can lead to errors in predictions of the model.

## 5. Conclusion

In this paper, we proposed a modified SEIR epidemic model for modeling COVID-19 in Iran and employed PSO algorithm to investigate the epidemic parameters and to predict the future trends of the epidemic.

The model estimated 15,252,372 people have been infected at the outbreak until April 9, 2021. Only 2,029,412 persons are officially confirmed. The results also indicated the probability of the fourth wave of COVID-19 cases, which can be the consequence of travelling and other events during Nowruz. Unless a strict social distancing is implemented immediately, the fourth wave of COVID-19 poses a tremendous burden on the health care system of Iran.

## Data Availability

The data that support the findings of this study are available from the corresponding author upon reasonable request. The data were derived from the following public domain resource: https://behdasht.gov.ir/

## Authors’ contributions

Ebrahim Sahafizadeh planned the experiments and wrote the manuscript; and MohammadAli Khajeian implemented the algorithms and conducted the experiments. All authors read and approved the final manuscript.

## References

[1] W. O. Kermack and a. G. McKendrick, “A Contributions to the mathematical theory of epidemics,” Proc. R. Soc. London, vol. 115, no. 772, pp. 700–721, 1927.

[2] Worldometer, “Iran Demographics 2020 (Population, Age, Sex, Trends),” Worldometer, 2020. [Online]. Available: https://www.worldometers.info/demographics/iran-demographics/. [Accessed: 10-Apr-2021].

[3] “Ministry of Health and Medical Education (MOHME).”.

[4] M. Clerc, “Particle Swarm Optimization,” Part. Swarm Optim., pp. 1942–1948, 2010.

[5] A. Annunziato and T. Asikainen, “Effective Reproduction Number Estimation from Data Series,” 2020. [Online]. Available: https://publications.jrc.ec.europa.eu/repository/bitstream/JRC121343/r0_technical_note_v3.4.pdf.

[6] ROBERT KOCH INSTITUT, “Epidemiologisches Bulletin,” 2020. [Online]. Available: https://www.rki.de/DE/Content/Infekt/EpidBull/Archiv/2020/Ausgaben/17_20.pdf?blob=publicationFile.

[7] E. Sahafizadeh and S. T. Azad, “365 days with COVID-19 in Iran: data analysis and epidemic curves,” medRxiv, pp. 1–5, 2021.

[8] A. Raoofi, A. Takian, A. Akbari Sari, A. Olyaeemanesh, H. Haghighi, and M. Aarabi, “COVID-19 Pandemic and Comparative Health Policy Learning in Iran,” Arch. Iran. Med., vol. 23, no. 4, pp. 220–234, 2020.

[9] A. Zahiri, S. Rafieenasab, and E. Roohi, “Prediction of Peak and Termination of Novel Coronavirus Covid-19 Epidemic in Iran,” vol. 98, no. 51, pp. 1–13, 2020.

[10] M. Moghadami, M. Hassanzadeh, K. Wa, A. Hedayati, and M. Malekolkalami, “Modeling the corona virus outbreak in IRAN,” medRxiv, 2020.

[11] E. Sahafizadeh and S. Sartoli, “Rising summer temperatures do not reduce the reproduction number of COVID-19,” J. Travel Med., vol. 2020, pp. 1–3, Oct. 2020.

[12] M. Ahmadi, A. Sharifi, S. Dorosti, S. Jafarzadeh Ghoushchi, and N. Ghanbari, “Investigation of effective climatology parameters on COVID-19 outbreak in Iran,” Sci. Total Environ., vol. 729, 2020.

[13] E. Sahafizadeh and S. Sartoli, “Epidemic curve and reproduction number of COVID-19 in Iran,” Journal of travel medicine, vol. 27, no. 5. 2020.

[14] N. Ghaffarzadegan and H. Rahmandad, “Simulation-based estimation of the spread of COVID-19 in Iran,” medRxiv. pp. 1–19, 2020.

[15] A. Ahmadi, M. Shirani, and F. Rahmani, “Modeling and Forecasting Trend of COVID-19 Epidemic in Iran,” medRxiv, p. 2020.03.17.20037671, 2020.

